# Molecular epidemiology of Western equine encephalitis virus in Brazil, 2023-2024

**DOI:** 10.1101/2024.04.15.24305848

**Authors:** Aline Scarpellini Campos, Ana Claúdia Franco, Fernanda Godinho, Rosana Huff, Jader da Cruz Cardoso, Paola Morais, Carolina Franceschina, Thales de Lima Bermann, Franciellen Machado dos Santos, Milena Bauermann, Tainá Machado Selayaran, Amanda Pellenz Ruivo, Cristiane Santin, Juciane Bonella, Carla Rodenbusch, José Carlos Ferreira, Scott C. Weaver, Vilar Ricardo Gewehr, Gabriel Luz Wallau, William M. de Souza, Richard Steiner Salvato

**Author notes:** These senior authors contributed equally to this work (William M. de Souza, Richard Steiner Salvato).

## Abstract

During the ongoing western equine encephalitis virus (WEEV) outbreak in South America, we described three fatal cases in horses from Rio Grande do Sul, Brazil. We sequenced WEEV strains and identified a novel lineage causing these cases. Continued surveillance and horse immunization are needed to mitigate the WEEV burden.

## Text

Western equine encephalitis virus (WEEV) is a mosquito-borne alphavirus that causes central nervous system (CNS) infection in humans and equids in the United States of America (USA), Canada, and the southern cone of South America (1). WEEV is transmitted in enzootic and epizootic transmission cycles mainly by mosquitoes of the *Culex* and *Aedes* genera among birds and lagomorphs, which can lead to sporadic spillover to equids and humans (2, 3). In humans, WEEV infections are usually mild or asymptomatic, with fever, headache, and myalgia (4). Some patients develop encephalitis, which can be fatal in 5%-15% of cases (5). In equids, WEEV infection can cause neurological disease (blindness, staggering, and seizures), often leading to death within days with high case-fatality rates. At present, no specific treatments or vaccines are available to treat or prevent WEEV infection in humans, but inactivated vaccines effectively prevent the disease in equines.

The largest WEEV outbreaks, with tens-of-thousands of equid and more than 3,000 human cases, were reported in the 1930s-40s. However, fewer than 700 confirmed cases were reported in the USA after the 1960s and none has been reported during the past 3 decades (1, 4). Similarly, major outbreaks occurred in South America during the 1970s and 1980s, followed by isolated cases mainly in Argentina in 1996 and Uruguay in 2009 (1, 6, 7). In December 2023, a significant WEEV re-emergence began with an ongoing major outbreak in Argentina and Uruguay. As of April 2024, there have been 103 confirmed human cases, 10 of them fatal, and 128 confirmed equine cases, with the majority fatal (8). In this study, we investigated active WEEV circulation in Rio Grande do Sul, Brazil, a state bordering Argentina, and Uruguay.

## The study

From December 2023 to April 2024, we conducted a two-pronged approach to investigate the presence of WEEV in Rio Grande do Sul, Brazil. First, we performed molecular screening in brain tissue samples from fatal horse cases, which were submitted to the Center for Health Surveillance of Rio Grande do Sul State between January 1, 2023 and April 10, 2024. Second, we carried out entomological surveillance in the Uruguaiana municipality. We focused on two horse breeding farms with a recent history of neurological equine disease, one case which was later confirmed as a WEEV-positive. Mosquitoes were collected using CDC light and Biogents BG-Pro traps placed near horses and vegetation, then classified based on morphological identification and partial sequencing of the *cytochrome oxidase I* gene (**Appendix**).

We captured 971 mosquitoes across seven genera that were combined into 117 pools for analysis. The most prevalent genus was *Culex*, with 68.4% (664/971). From January 1, 2023 and April 10, 2024, we received brain samples of 31 fatal horse cases from 4.4% (22/497) of municipalities in Rio Grande do Sul State. Brain tissue fragments (≈ 2 g/cm^3^) were homogenized with 800μL of TRIzol, while mosquitoes were homogenized with phosphate-buffered saline. Next, we extracted RNA from all samples and performed real-time reverse transcription-PCR (rRT-PCR) to detect WEEV RNA (9). Then, we carried out sequencing using the Illumina Viral Surveillance Panel (Illumina, USA) and conducted maximum-likelihood phylogenetic inferences **(Appendix)**.

Of 31 horse brain tissue samples tested by rRT-PCR, we detected WEEV RNA in 3 (9.7%) with cycle threshold (Ct) values of 26-27 (**Fig. A**). All three horses exhibited signs of neurological disease (i.e., paralysis and incoordination) and were not vaccinated against WEEV. The first two horses were two months and two years old, respectively, and died on December 21 and 28, 2023, in Barra do Quaraí and Uruguaiana municipalities, respectively. These municipalities are located on the Brazilian border with Argentina and Uruguay. The third case was a five-month-old horse that died on January 30, 2024, in Jaguarão municipality, bordering Uruguay. Additionally, 3 fatal horse cases were positive for Rabies virus by rRT-PCR **(Appendix Table S1)**. All mosquito pools tested negative for WEEV, eastern equine encephalitis, West Nile, St. Louis encephalitis, Mayaro, and Oropouche viruses.

**Figure 1.**
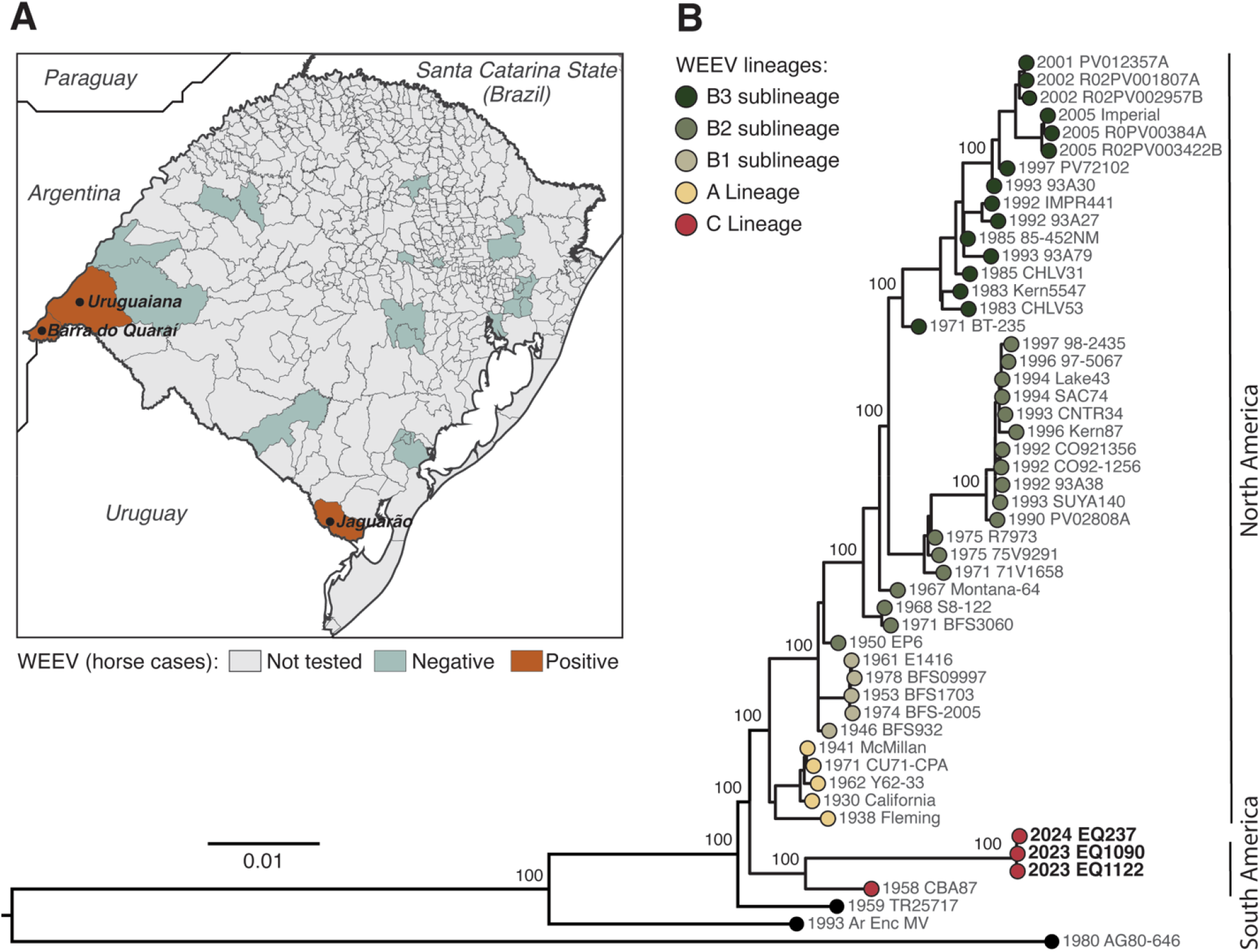
Molecular epidemiology of Western equine encephalitis virus in Rio Grande do Sul State, Brazil. **A**) Map of fatal horse cases positive by rRT-PCR for WEEV (red), per municipality, in Rio Grande do Sul State between December 2023 and April 2024. The cases occurred occurred in Barra do Quaraí on 21 December 2023 (EQ1090), Uruguaiana on 28 December 2023 (EQ1122), and in Jaguarão on 30 January 2024 (EQ237). The municipalities tested are described in Appendix Table S1. **B**) Maximum likelihood phylogenetic tree of WEEV, including three new WEEV genomes (red dots) from Rio Grande do Sul State generated in this study. The phylogeny is midpoint rooted. The scale bar indicates the evolutionary distance of substitutions per nucleotide. Bootstrap values based on 1,000 replicates are shown on principal nodes. The GenBank accession numbers of sequences used in this figure are presented in Appendix Table S4.

Next, we used Illumina sequencing to generate the nearly complete (≥98%) coding sequences for three WEEV strains with a mean depth of coverage of 606-fold/nucleotide. We submitted sequences to GenBank (accession nos. PP544260, XXXXXXX, XXXXXXX). The maximum-likelihood phylogenetic analysis showed that three WEEV strains circulating in the Rio Grande do Sul State in 2024 clustered together in a distinct, well-supported clade (100% bootstrap). This clade represents a novel WEEV lineage closely related to the CB87 strain documented in the 1960s in Argentina, which we proposed as the C lineage (**Fig. B**). These novel strains shared 97-100% nucleotide sequence identity, including with the CB87 strain. No evidence of recombination was found in these WEEV strains.

## Conclusion

We identified a novel WEEV lineage associated with the ongoing outbreak in South America, detected in three fatal horse cases from Rio Grande do Sul, Brazil. This new lineage forms a distinct evolutionary clade evolving independently for many decades from those circulating in North America (1, 4, 10). These findings suggest that WEEV has been circulating in South America over the past 15 years, with cases likely unreported. We hypothesize two possible explanations for this recent lack of reported WEEV cases. Limited enzootic circulation of the WEEV between vectors and resident/migratory birds, or a lack of WEEV surveillance, particularly in rural or remote areas, where cases might go undetected.

Some experimental murine model studies indicate reduced virulence of recent WEEV isolates (B3 strains) compared to earlier isolates (A, B1, B2 strains), which could explain decreased human and equine cases in North America (1, 4, 11). However, the large numbers of neurological and fatal cases in humans and equines in Argentina, Brazil, and Uruguay (2023-2024), suggest that contemporary WEEV strains circulating in South America might be more virulent than those currently circulating in North America. Further research is needed to elucidate the virulence determinants that might explain this apparent difference between South and North American WEEV lineages.

This study has several limitations. First, we focused on identifying active WEEV infections using molecular methods only in fatal horse cases. Horses with mild signs of the disease should also be tested. Also, age-stratified serological studies are needed to determine the extent of previous WEEV exposure in the equine and human populations and patterns of past infections. Second, we did not detect the WEEV RNA in mosquitoes. Although our entomological investigation shows that the *Culex spp*. were the most abundant mosquito species in the region, further studies with an emphasis on the previously incriminated *Aedes albifasciatus* (3, 12) are needed to better understand the transmission dynamics of WEEV in South America. Third, we were unable to determine whether ecological drivers were associated with the current outbreaks, but further studies should investigate the climate factors, anthropogenic changes, and migratory bird routes and activity (13).

In conclusion, our study identified active WEEV circulation in Rio Grande do Sul, Brazil, and a novel viral lineage associated with fatal horse cases. These findings highlight the critical need for continuous laboratory diagnosis and surveillance for WEEV in both horses and humans, as well as ecological studies using a One Health approach to understand better the transmission dynamics and ecological drivers contributing to WEEV re-emergence in South America. Finally, horse immunization should be considered to mitigate the impact on animal health.

## Data Availability

All data produced in the present study are available upon reasonable request to the authors

