## Appendix for "Molecular epidemiology of Western equine encephalitis virus in Brazil, 2023-2024"

**Materials and Methods**

**Equine samples**

Brain samples from horses with neurological signs were collected during necropsies in 22 of 497(4.4%) municipalities of Rio Grande do Sul State, Brazil. The State Department of Agriculture collected the samples and then sent them to the Rio Grande do Sul State Center for Health Surveillance to investigate the etiological agent. All samples were stored at −80°C. The information on horse samples is presented in Appendix Table 1.

**Entomological surveillance**

We deployed 8 CDC light traps and 4 Biogents BG-Pro traps positioned at a height of 1.5 meters above ground level, placed near horses and vegetation. Mosquito sampling was conducted during two time periods at each site: a 24-hour period starting at 6 pm and a 12-hour period from 6 pm to 6 am. Captured mosquitoes were then flash-frozen, stored in liquid nitrogen, and transported for storage at -80°C at the Rio Grande do Sul State Center for Health Surveillance in Porto Alegre City. Finally, mosquito species were identified morphologically using standard keys (1, 2). Then, pools were formed by species, with a maximum of 10 mosquitoes per pool. The *cytochrome oxidase I* gene was additionally sequenced for the molecular identification of mosquito pools, following previously described methods (3). The information on mosquito pool samples are presented in Appendix Table 2.

**Real-time quantitative reverse transcription-polymerase chain reaction (rRT-PCR) for Western equine encephalitis virus, Eastern equine encephalitis, West Nile, St. Louis encephalitis, Mayaro, Oropouche, and rabies viruses**

Brain tissue fragments (≈ 2 g/cm³) were immersed in 800 µl of TRIzol reagent and subjected to disruption using a Precellys® 24 Touch (Thomas Scientific, USA). Subsequently, the mixture was immediately centrifuged to isolate the supernatant. Viral RNA was extracted from the supernatant using the Extracta Kit Fast–DNA and RNA Viral (Loccus, Brazil) following the manufacturer's instructions. Then, extracted RNA was tested by rRT-PCR targeting western equine encephalitis virus using the TaqMan RNA to-CT 1-Step Kit (Applied Biosystems, USA) as previously described elsewhere (4). Reactions were performed on CFX Opus 96 Real-Time PCR System (Bio-Rad, USA). Additionally, the mosquito pool samples were rested by rRT-PCR targeting eastern equine encephalitis (Madariaga) (4), West Nile and St. Louis encephalitis (5), Mayaro and Oropouche (6), and rabies viruses (7) as described in Appendix Table 3.

**Western equine encephalitis virus genome sequencing and assembly**

WEEV genome sequencing was conducted on three horse brain samples that tested positive by rRT-PCR. The near-complete genome was achieved using the hybrid–capture-based metagenomic approach enabled by the Illumina Viral Surveillance Panel (VSP) and Illumina RNA Prep with Enrichment kit (Cat no. 20087932), according to the manufacturer's instructions. VSP-enriched libraries were sequenced on an Illumina MiSeq platform. The generated raw FASTQ files were processed through the ViralFlow 1.0 pipeline (8) for assembly utilizing the WEEV strain 71V-1658 (GenBank accession no. NC_003908.1), as a reference genome.

**Phylogenetic analysis**

The three novel WEEV genomes with >90% coverage were generated and aligned with WEEV strains with complete coding sequences available in the GenBank database as of April 10, 2024. Then, we performed a multiple sequence alignment (MSA) built using MAFFT version 7.450 (9), and manual adjustment was conducted using Geneious Prime 2023.0.4. The dataset was screened for recombination events using all available methods in RDP version 4 (10), but no evidence of recombination was found. A maximum likelihood (ML) phylogeny tree was performed using IQ-TREE version 2 under a GTR + I + γ model determined by ModelFinder (11, 12). The ultrafast-bootstrap approach with 1,000 replicates was used to determine the statistical support for nodes for the ML phylogeny (13). The phylogenetic tree was visualized using Figtree 1.4.2 (http://tree.bio.ed.ac.uk/software/figtree/).

**Supplementary Tables**

**Table S1.** Characteristics of samples of horses exhibiting neurological signs submitted to the Rio Grande do Sul State Center for Health Surveillance.

| **Municipality** | **Date of Collection** | **Age** | **WEEV** | **Rabies -**  **Ct-value** |
| --- | --- | --- | --- | --- |
| Morro Redondo | 24-Jan-2023 | 11 years | ND | ND |
| Pelotas | 24-Jan-2023 | 15 years | ND | ND |
| Gravataí | 17-Feb-2023 | 14 years | ND | ND |
| Taquara | 4-Apr-2023 | 4 years | ND | ND |
| Parobé | 10-Apr-2023 | 2 years | ND | ND |
| Pantano Grande | 2-May-2023 | 8 years | ND | ND |
| Rio Pardo | 10-May-2023 | 12 years | ND | Positive |
| Alegrete | 10-May-2023 | 5 years | ND | ND |
| Porto Alegre | 30-May-2023 | N/A | ND | ND |
| Porto Alegre | 30-May-2023 | N/A | ND | ND |
| Igrejinha | 20-June-2023 | 4 years | ND | ND |
| Porto Alegre | 21-June-2023 | 11 years | ND | ND |
| Porto Alegre | 13-July-2023 | 12 years | ND | ND |
| Glorinha | 28-July-2023 | N/A | ND | ND |
| Porto Alegre | 22-Aug-2023 | 23 years | ND | ND |
| Bossoroca | 24-Aug-2023 | 5 years | ND | ND |
| Marau | 19-Sep-2023 | 8 years | ND | ND |
| Boqueirão do Leão | 28-Nov-2023 | N/A | ND | ND |
| Boqueirão do Leão | 28-Nov-2023 | N/A | ND | ND |
| Arroio do Meio | 13-Dec-2023 | 12 years | ND | ND |
| Barra do Quaraí | 21-Dec-2023 | 2 months | Positive (Ct-value 26) | ND |
| São Miguel das Missões | 27-Dec-2023 | 4 years | ND | Positive |
| Caxias do Sul | 27-Dec-2023 | 14 years | ND | ND |
| Bagé | 28-Dec-2023 | 2 months | ND | ND |
| Uruguaiana | 28-Dec-2023 | 2 years | Positive (Ct-value 27) | ND |
| Alegrete | 5-Jan-2024 | N/A | ND | Positive |
| Jaguarão | 30-Jan-2024 | 5 months | Positive (Ct-value 27) | ND |
| Porto Alegre | 5-Feb-2024 | N/A | ND | ND |
| Porto Alegre | 16-Feb-2024 | 12 years | ND | ND |
| Itaqui | 21-Feb-2024 | 14 years | ND | ND |
| Gravataí | 6-Mar-2024 | N/A | ND | ND |

Legend: Ct-value, cycle threshold value. WEEV, Western equine encephalitis virus**.** N/A, unavailable. ND, Not detected.

**Table S2.** Characteristics of mosquitoes collecting during the entomological surveillance in Uruguaiana municipality, Rio Grande do Sul State, Brazil.

| **ID** | **Pool size** | **Species** |
| --- | --- | --- |
| MQ1.1 | 1 | *Aedeomyia squamipennis* |
| MQ1.2 | 1 | *Mansonia sp.* |
| MQ1.3 | 10 | *Culex sp.* |
| MQ1.4 | 4 | *Culex sp.* |
| MQ10 | 1 | *Culex sp.* |
| MQ11 | 2 | *Culex pipiens* |
| MQ12.1 | 1 | *Culex coronator* |
| MQ12.2 | 1 | *Culex maxi* |
| MQ2.1 | 10 | *Culex pipiens* |
| MQ2.10 | 10 | *Culex pipiens* |
| MQ2.11 | 10 | *Anopheles deaneorum* |
| MQ2.12 | 10 | *Anopheles albitarsis* |
| MQ2.13 | 10 | *Mansonia titillans* |
| MQ2.14 | 10 | *Mansonia titillans* |
| MQ2.15 | 10 | *Culex coronator* |
| MQ2.16 | 10 | *Culex coronator* |
| MQ2.17 | 10 | *Culex maxi* |
| MQ2.18 | 10 | *Uranotaenia lowii* |
| MQ2.19 | 10 | *Culex coronator* |
| MQ2.2 | 10 | *Culex coronator* |
| MQ2.20 | 10 | *Culex pipiens* |
| MQ2.21 | 10 | *Mansonia titillans* |
| MQ2.22 | 10 | *Culex coronator* |
| MQ2.23 | 10 | *Culex coronator* |
| MQ2.24 | 10 | *Mansonia titillans* |
| MQ2.25 | 10 | *Anopheles deaneorum* |
| MQ2.26 | 10 | *Culex coronator* |
| MQ2.27 | 10 | *Culex coronator* |
| MQ2.28 | 10 | *Culex coronator* |
| MQ2.29 | 10 | *Culex interfor* |
| MQ2.3 | 10 | *Culex pipiens* |
| MQ2.30 | 10 | *Uranotaenia lowii* |
| MQ2.31 | 10 | *Culex pipiens* |
| MQ2.32 | 10 | *Culex pipiens* |
| MQ2.33 | 10 | *Culex coronator* |
| MQ2.34 | 10 | *Culex pipiens* |
| MQ2.35 | 10 | *Culex pipiens* |
| MQ2.36 | 10 | *Anopheles albitarsis* |
| MQ2.37 | 10 | *Anopheles albitarsis* |
| MQ2.38 | 5 | *Aedeomyia squamipennis* |
| MQ2.39 | 5 | *Culex coronator* |
| MQ2.4 | 10 | *Culex coronator* |
| MQ2.40 | 3 | *Culex intrincatus* |
| MQ2.41 | 1 | *Psorophora confinnis* |
| MQ2.42 | 2 | *Aedes crinifer* |
| MQ2.43 | 1 | *Culex coronator* |
| MQ2.5 | 10 | *Culex coronator* |
| MQ2.6 | 10 | *Mansonia sp.* |
| MQ2.7 | 10 | *Culex pipiens* |
| MQ2.8 | 10 | *Culex pipiens* |
| MQ2.9 | 10 | *Culex sp.* |
| MQ3.1 | 10 | *Culex pipiens* |
| MQ3.2 | 5 | *Culex pipiens* |
| MQ4.1 | 10 | *Culex coronator* |
| MQ4.10 | 10 | *Culex coronator* |
| MQ4.11 | 10 | *Culex pipiens* |
| MQ4.12 | 10 | *Anopheles albitarsis* |
| MQ4.13 | 10 | *Anopheles albitarsis* |
| MQ4.14 | 10 | *Uranotaenia lowii* |
| MQ4.15 | 10 | *Culex pipiens* |
| MQ4.16 | 10 | *Culex pipiens* |
| MQ4.17 | 10 | *Culex coronator* |
| MQ4.18 | 10 | *Anopheles deaneorum* |
| MQ4.19 | 10 | *Culex coronator* |
| MQ4.2 | 10 | *Culex interfor* |
| MQ4.20 | 10 | *Culex pipiens* |
| MQ4.21 | 10 | *Uranotaenia lowii* |
| MQ4.22 | 10 | *Anopheles deaneorum* |
| MQ4.23 | 10 | *Culex coronator* |
| MQ4.24 | 10 | *Culex pipiens* |
| MQ4.25 | 10 | *Uranotaenia lowii* |
| MQ4.26 | 10 | *Anopheles albitarsis* |
| MQ4.27 | 10 | *Culex coronator* |
| MQ4.28 | 10 | *Culex coronator* |
| MQ4.29 | 10 | *Culex coronator* |
| MQ4.3 | 10 | *Culex sp.* |
| MQ4.30 | 10 | *Anopheles deaneorum* |
| MQ4.31 | 10 | *Culex coronator* |
| MQ4.32 | 6 | *Culex maxi* |
| MQ4.33 | 4 | *Psorophora confinnis* |
| MQ4.34 | 4 | *Psorophora confinnis* |
| MQ4.4 | 10 | *Anopheles albitarsis* |
| MQ4.5 | 10 | *Uranotaenia lowii* |
| MQ4.6 | 10 | *Culex pipiens* |
| MQ4.7 | 10 | *Culex coronator* |
| MQ4.8 | 10 | *Anopheles deaneorum* |
| MQ4.9 | 2 | *Aedes scapularis* |
| MQ5 | 1 | *Culex coronator* |
| MQ6 | 6 | *Culex pipiens pallens* |
| MQ7.1 | 10 | *Culex pipiens* |
| MQ7.10 | 10 | *Culex pipiens* |
| MQ7.11 | 5 | *Anopheles albitarsis* |
| MQ7.12 | 1 | *Aedes scapularis* |
| MQ7.13 | 2 | *Ochlerotatus scapularis* |
| MQ7.14 | 2 | *Aedes aegypti* |
| MQ7.2 | 10 | *Culex pipiens* |
| MQ7.3 | 10 | *Culex pipiens* |
| MQ7.4 | 10 | *Anopheles darlingi* |
| MQ7.5 | 10 | *Anopheles sp.* |
| MQ7.6 | 6 | *Aedes scapularis* |
| MQ7.7 | 10 | *Culex pipiens* |
| MQ7.8 | 10 | *Culex pipiens* |
| MQ7.9 | 4 | *Culex pipiens* |
| MQ8.1 | 10 | *Culex pipiens* |
| MQ8.10 | 10 | *Culex pipiens* |
| MQ8.11 | 10 | *Culex pipiens* |
| MQ8.12 | 10 | *Culex pipiens* |
| MQ8.13 | 10 | *Culex pipiens* |
| MQ8.14 | 7 | *Culex bidens* |
| MQ8.2 | 10 | *Culex pipiens* |
| MQ8.3 | 10 | *Culex pipiens* |
| MQ8.4 | 7 | *Culex pipiens* |
| MQ8.5 | 1 | *Anopheles albitarsis* |
| MQ8.6 | 10 | *Aedes albopictus* |
| MQ8.7 | 10 | *Culex pipiens* |
| MQ8.8 | 10 | *Culex pipiens* |
| MQ8.9 | 10 | *Culex pipiens* |

**Table S3.** Primers and probes were used for the detection of viruses in this study.

| **Virus** | **Sequences (5’→3’)** | **Primers and probes** | **Target** | **Ref.** |
| --- | --- | --- | --- | --- |
| RABV | ACGCTTAACAACCAGATCAAAGAA | Forward | LN34 | (Wadhwa et al., 2017) |
|  | ACGCTTAACAACAAAATCADAGAAG | Forward | LN34 |  |
|  | CMGGGTAYTTRTAYTCATAYTGRTC | Reverse | N34 |  |
|  | FAM-AACACCYCTACAATGGA-BHQ1 | Probe | N34 |  |
|  | FAM AACACTACTACAATGGA-BHQ1 | Probe | N34 |  |
|  | CGATGAAGATCAAGATCATTGC | Forward | β-Actin |  |
|  | AAGCATTTGCGGTGGAC | Reverse | β-Actin |  |
|  | HEX-TCCACCTTCCAGCAGATGTGGATCA-BHQ1 | Probe | β-Actin |  |
| MAYV | CACGGACMTTTTGCCTTCA | Forward | NSP1 | Naveca et al., 2017 |
|  | AGACTGCCACCTCTGCTKGAG | Reverse | NSP1 |  |
|  | VIC-ACAGATCAGACATGCAGG | Probe | NSP1 |  |
| OROV | TCCGGAGGCAGCATATGTG | Forward | NP | Naveca et al., 2017 |
|  | ACAACACCAGCATTGAGCACTT | Reverse | NP |  |
|  | FAM-CATTTGAAGCTAGATACGG | Probe | NP |  |
| WNV | CAGACCACGCTACGGCG | Forward | E | Lanciotti et al., 2001 |
|  | CTAGGGCCGCGTGGG | Reverse | E |  |
|  | TCTGCGGAGAGTGCAGTCTGCGAT | Probe | E |  |
| SLEV | CTGGCTGTCGGAGGGATTCT | Forward | E | Lanciotti et al., 2001 |
|  | TAGGTCAATTGCACATCCCG | Reverse | E |  |
|  | TCTGGCGACCAGCGTGCAAGCCG | Probe | E |  |
| EEEV | ACACCGCACCCTGATTTTACA | Forward | E2 | Lambert et al., 2001 |
|  | CTTCCAAGTGACCTGGTCGTC | Reverse | E2 |  |
|  | TGCACCCGGACCATCCGACCT | Probe | E2 |  |
| WEEV | CTGAAAGTCGGCCTGCGTAT | Forward | E2 | Lambert et al., 2001 |
|  | CGCCATTGACGAACGTATCC | Reverse | E2 |  |
|  | ATACGGCAATACCACCGCGCACC | Probe | E2 |  |

Legend: RABV, Rabies virus. MAYV, Mayaro virus. OROV, Oropouche virus. WNV, West Nile virus. SLEV, St Louis encephalitis virus. WEEV, Western equine encephalitis virus

**Table S4.** Genome sequences used in the phylogenetic analysis.

| **Accession** | **Isolate** | **Location** | **Host** | **Year** | **Lineage** |
| --- | --- | --- | --- | --- | --- |
| GQ287640 | McMillan | Canada: Ontario | Homo sapiens | 1941 | A |
| KJ554965 | California | USA: San Joaquin Valley, CA | Equus caballus | 1930 | A |
| KT844544 | Y62-33 | Russia: Urdmurt | Aedes cinereus | 1962 | A |
| KT844545 | CU71-CPA | Cuba |  | 1971 | A |
| MN477208 | Fleming | USA | Homo sapiens | 1938 | A |
| GQ287644 | BFS-2005 | USA: Kern County, California | Culex tarsalis | 1974 | B1 |
| KJ554966 | BFS932 | USA: Bakersfield, CA | Culex tarsalis | 1946 | B1 |
| KJ554968 | BFS1703 | USA: Bakersfield, CA | Culex tarsalis | 1953 | B1 |
| KJ554969 | E1416 | USA: Kern Co., CA | Zonotrichia leucophrys | 1961 | B1 |
| KJ554974 | BFS09997 | USA: Kern Co., CA | Culex tarsalis | 1978 | B1 |
| KJ554973 | 75V9291 | USA: Wilkin City, MN | Culex tarsalis | 1975 | B2 |
| KJ554977 | PV02808A | USA: Lubbock Co., TX | Culicidae | 1990 | B2 |
| KJ554979 | CO921356 | USA: Larimer City, CO | Culex tarsalis | 1992 | B2 |
| KJ554980 | 93A38 | USA: Tacna, AZ | Culicidae | 1992 | B2 |
| KJ554984 | CNTR34 | USA: Contra Costa Co., CA | Culex tarsalis | 1993 | B2 |
| KJ554985 | Lake43 | USA: Lake Co., CA | Culex tarsalis | 1994 | B2 |
| KT844546 | CO92-1256 | USA: Colorado, Larimer City | Culex tarsalis | 1992 | B2 |
| KT844548 | SUYA140 | USA: California, Sutter County | Culex tarsalis | 1993 | B2 |
| KT844549 | SAC74 | USA: California, Sacramento County | Culex tarsalis | 1994 | B2 |
| KT844550 | Kern87 | USA: California, Kern County | Culex tarsalis | 1996 | B2 |
| KU978771 | 97-5067 | USA | Meleagris gallopavo | 1996 | B2 |
| KU978772 | 98-2435 | USA | Dromaius novaehollandiae | 1997 | B2 |
| NC_003908 | 71V1658 | USA: Oregon |  | 1971 | B2 |
| OQ184867 | R7973 | USA: Larimer County, Colorado | Homo sapiens | 1975 | B2 |
| KJ554967 | EP6 | USA: Missouri | Culicidae | 1950 | B2 |
| GQ287641 | Imperial | USA: Imperial County, California | Culex tarsalis | 2005 | B3 |
| GQ287643 | Montana-64 | USA: Montana | Equus caballus | 1967 | B3 |
| GQ287647 | 85-452NM | USA: New Mexico | Culex tarsalis | 1985 | B3 |
| KJ554970 | S8-122 | USA: Butte Co., CA | Hesperosciurus griseus | 1968 | B3 |
| KJ554971 | BT-235 | USA: Texas | Gopherus berlandieri | 1971 | B3 |
| KJ554972 | BFS3060 | USA: Butte Co., CA | Culex tarsalis | 1971 | B3 |
| KJ554975 | KERN5547 | USA: Kern Co., CA | Culex tarsalis | 1983 | B3 |
| KJ554976 | CHLV53 | USA: Riverside Co., CA | Culex tarsalis | 1983 | B3 |
| KJ554978 | IMPR441 | USA: Imperial Co., CA | Culex tarsalis | 1992 | B3 |
| KJ554981 | 93A27 | USA: Parker, AZ | Culicidae | 1992 | B3 |
| KJ554982 | 93A30 | USA: Phoenix, AZ | Culicidae | 1993 | B3 |
| KJ554983 | 93A79 | USA: Yuma, AZ | Culicidae | 1993 | B3 |
| KJ554986 | PV72102 | USA: El Paso Co., TX | Culicidae | 1997 | B3 |
| KJ554987 | PV012357A | USA: El Paso Co., TX | Culicidae | 2001 | B3 |
| KJ554988 | R02PV002957B | USA: El Paso Co., TX | Culicidae | 2002 | B3 |
| KJ554989 | R02PV001807A | USA: El Paso Co., TX | Culicidae | 2002 | B3 |
| KJ554990 | R02PV003422B | USA: El Paso Co., TX | Culicidae | 2005 | B3 |
| KJ554991 | R0PV00384A | USA: El Paso Co., TX | Culicidae | 2005 | B3 |
| KT844547 | CHLV31 | USA: California, Riverside County | Culex tarsalis | 1985 | B3 |
| KT844543 | CBA87 | Argentina: Cordoba Pr., Oncativo | Equus ferus | 1958 | C |
| PP544260 | EQ1090 | Brazil: Rio Grande do Sul | Equus caballus | 2023 | C |
| To be release | EQ1122 | Brazil: Rio Grande do Sul | Equus caballus | 2023 | C |
| To be release | EQ237 | Brazil: Rio Grande do Sul | Equus caballus | 2024 | C |
| KT844541 | TR25717 | Guyana | Equus ferus | 1959 | none |
| KT844542 | Ar_Enc_MV | Argentina: Buenos Aires Pr., Monte Veloz | Equus ferus | 1993 | none |
| NC_075015 | AG80-646 | Argentina: Chaco Province | Culex | 1980 | none |
